# Neutrophil extracellular traps and thrombosis in COVID-19

**DOI:** 10.1101/2020.04.30.20086736

**Authors:** Yu Zuo, Melanie Zuo, Srilakshmi Yalavarthi, Kelsey Gockman, Jacqueline A. Madison, Hui Shi, Wrenn Woodward, Sean P. Lezak, Njira L. Lugogo, Jason S. Knight, Yogendra Kanthi

## Abstract

**Background:** Early studies of patients with COVID-19 have demonstrated markedly dysregulated coagulation and a high risk of morbid arterial and venous thrombotic events. While elevated levels of blood neutrophils and neutrophil extracellular traps (NETs) have been described in patients with COVID-19, their potential role in COVID-19-associated thrombosis remains unknown.

**Objectives:** To elucidate the potential role of hyperactive neutrophils and NET release in COVID-19-associated thrombosis.

**Patients/Methods:** This is a retrospective, case-control study of patients hospitalized with COVID-19 who developed thrombosis (n=11), as compared with gender- and age-matched COVID-19 patients without clinical thrombosis (n=33). In addition to capturing clinical data, we measured remnants of NETs (cell-free DNA, myeloperoxidase-DNA complexes, and citrullinated histone H3) and neutrophil-derived S100A8/A9 (calprotectin) in patient sera.

**Results:** The majority of patients (9/11) were receiving at least prophylactic doses of heparinoids at the time thrombosis was diagnosed. As compared with controls, patients with COVID-19-associated thrombosis had significantly higher blood levels of markers of NETs (cell-free DNA, myeloperoxidase-DNA complexes, citrullinated histone H3) and neutrophil activation (calprotectin). The thrombosis group also had higher levels of D-dimer, CRP, ferritin, and platelets, but not troponin or neutrophils. Finally, there were strong associations between markers of hyperactive neutrophils (calprotectin and cell-free DNA) and D-dimer.

**Conclusion:** Elevated levels of neutrophil activation and NET formation in patients hospitalized with COVID-19 are associated with higher risk of morbid thrombotic complications. These observations underscore the need for urgent investigation into the potential relationship between NETs and unrelenting thrombosis in COVID-19.

**ESSENTIALS:**

- Mechanisms contributing to frequent thrombosis in COVID-19 remain unknown
- NETs and neutrophil activation were measured in patients with COVID-19 -associated thrombosis
- Thrombosis in COVID-19 was associated with higher levels of circulating NETs and calprotectin
- Contributions of neutrophils and NETs to thrombosis in COVID-19 warrant urgent investigation

## INTRODUCTION

Severe acute respiratory syndrome coronavirus 2 (**SARS-CoV-2**) causes the disease known as coronavirus disease 2019 (**COVID-19**). It most commonly presents with influenza-like illness and viral pneumonia, but in its most severe manifestation progresses to acute respiratory distress syndrome (**ARDS**) and multi-organ failure [1]. To date the viral pandemic has resulted in millions of infections worldwide (https://coronavirus.jhu.edu/map.html).

In COVID-19, elevated levels of blood neutrophils predict severe respiratory disease and unfavorable outcomes [2, 3]. Neutrophil-derived neutrophil extracellular traps (**NETs**) play a pathogenic role in many thrombo-inflammatory states including sepsis [4, 5], thrombosis [6-8], and respiratory failure [9, 10]. NETs are extracellular webs of chromatin and microbicidal proteins that are an evolutionarily conserved aspect of innate immune host-defense [11]; however, NETs also have potential to initiate and propagate inflammation and thrombosis. NETs deliver a variety of oxidant enzymes to the extracellular space, including myeloperoxidase, NADPH oxidase, and nitric oxide synthase, while also serving as a source of extracellular histones that carry significant cytotoxic potential. NETs drive cardiovascular disease by propagating inflammation in vessel walls [12]. Furthermore, when formed intravascularly, NETs can occlude arteries [13], veins [14], and microscopic vessels [15]. Early studies of COVID-19 suggest a high risk of morbid arterial events [16], and the risk of venous thromboembolism (**VTE**) is increasingly revealing itself as more data become available [17].

Descriptive and mechanistic studies to date that examine COVID-19 pathophysiology have focused on monocytes and lymphocytes more so than neutrophils and their effector products. Our group recently reported high levels of NETs in 50 patients hospitalized with COVID-19 as compared with healthy controls [18]. Here, we describe 11 cases of thrombosis in patients hospitalized with COVID-19 and demonstrate an association with neutrophil hyperactivity and NET release.

## METHODS

### Human samples for NETs analysis

All 44 patients studied here had a confirmed COVID-19 diagnosis based on FDA-approved DiaSorin Molecular Simplexa COVID-19 Direct real-time RT-PCR assay. Blood was collected into serum separator tubes by a trained hospital phlebotomist. After completion of biochemical testing ordered by the clinician, the remaining serum was stored at 4°C for 4 to 48 hours before it was deemed "discarded” and released to the research laboratory. Serum samples were immediately divided into small aliquots and stored at -80°C until the time of testing. This study complied with all relevant ethical regulations, and was approved by the University of Michigan Institutional Review Board (HUM00179409), which waived the requirement for informed consent given the discarded nature of the samples.

### Quantification of S100A8/A9 (calprotectin)

Calprotectin levels were measured with the Human S100A8/S100A9 Heterodimer DuoSet ELISA (DY8226-05, R&D Systems) according to the manufacturer’s instructions.

### Quantification of cell-free DNA

Cell-free DNA was quantified in sera using the Quant-iT PicoGreen dsDNA Assay Kit (Invitrogen, P11496) according to the manufacturer’s instructions.

### Quantification of myeloperoxidase-DNA complexes

Myeloperoxidase-DNA complexes were quantified similarly to what has been previously described [19]. This protocol used several reagents from the Cell Death Detection ELISA kit (Roche). First, a high-binding EIA/RIA 96-well plate (Costar) was coated overnight at 4°C with anti-human myeloperoxidase antibody (Bio-Rad 0400-0002), diluted to a concentration of 1 µg/ml in coating buffer (Cell Death kit). The plate was washed two times with wash buffer (0.05% Tween 20 in PBS), and then blocked with 4% bovine serum albumin in PBS (supplemented with 0.05% Tween 20) for 2 hours at room temperature. The plate was again washed five times, before incubating for 90 minutes at room temperature with 10% serum or plasma in the aforementioned blocking buffer (without Tween 20). The plate was washed five times, and then incubated for 90 minutes at room temperature with 10x anti-DNA antibody (HRP-conjugated; from the Cell Death kit) diluted 1:100 in blocking buffer. After five more washes, the plate was developed with 3,3‘,5,5’-Tetramethylbenzidine (TMB) substrate (Invitrogen) followed by a 2N sulfuric acid stop solution. Absorbance was measured at a wavelength of 450 nm using a Cytation 5 Cell Imaging Multi-Mode Reader (BioTek). Data were normalized to *in vitro-prepared* NET standards included on every plate, which were quantified based on their DNA content.

### Quantification of citrullinated-histone H3

Citrullinated-histone H3 was quantified in sera using the Citrullinated Histone H3 (Clone 11D3) ELISA Kit (Cayman, 501620) according to the manufacturer’s instructions.

### Statistical analysis

When two groups were present, data were analyzed by Mann-Whitney test. Correlations were tested by Pearson’s correlation coefficient. Data analysis was with GraphPad Prism software version 8. Statistical significance was defined as p<0.05.

## RESULTS

We identified 11 patients who developed a thrombotic event while admitted for treatment of COVID-19 at a large academic medical center (**Table 1**). The events consisted of three strokes and nine VTE events (**Table 2**). Nine were receiving at least prophylactic doses of heparinoids at the time the event was diagnosed. The other two patients were found to have pulmonary emboli on the day of admission (**Table 2**). Nine of the patients were receiving respiratory support by mechanical ventilation at the time of the event. To date, eight have been discharged from the hospital, two have died, and one remains hospitalized.

**Table 1:** COVID-19 patient characteristics.

| Table 1: COVID-19 patient characteristics |  |  |
| --- | --- | --- |
|  | Thrombosis (n=11) | Matched (n=33) |
| **Demographics** |  |  |
| Age (years)* | 56 ± 12 (38-77) | 57 ± 12 (33-82) |
| Female | 2 (18.1%) | 6 (18.2%) |
| White/Caucasian | 3 (27.2%) | 14 (42.4%) |
| Black/African-American | 5 (45.5%) | 15 (45.5%) |
| Unknown | 2 (18.2%) | 4 (12.1%) |
| **Thrombosis** |  |  |
| Arterial | 2 (18.1%) | 0 (0%) |
| Venous | 8 (72.7%) | 0 (0%) |
| Both | 1 (9%) | 0 (0%) |
| **Comorbidities** |  |  |
| Ischemic heart disease | 5 (45.5%) | 8 (24.2%) |
| History of stroke | 1 (9%) | 4 (12.1%) |
| Hypertension | 6 (54.5%) | 22 (66.7%) |
| Obesity | 6 (54.5%) | 22 (66.7%) |
| History of smoking | 4 (36.4%) | 8 (24.2%) |
| Diabetes | 4 (36.4%) | 15 (45.5%) |
| Renal disease | 4 (36.4%) | 10 (30.3%) |
| Lung disease | 1 (9%) | 6 (18.2%) |
| Cancer | 1 (9%) | 6 (18.2%) |
| Autoimmune disease | 1 (9%) | 0 (0%) |
| Immune deficiency | 0 | 4 (12.1%) |
| **Outcome** |  |  |
| Discharged | 8 (72.7%) | 23 (69.5%) |
| Death | 2 (18.1%) | 7 (21.2%) |
| Remains hospitalized | 1 (9%) | 3 (9%) |
| * Mean ± standard deviation |  |  |

**Table 2:** Thrombosis details in patients with COVID-19.

| Patient | Age | Sex | Day | Ventilation | Event | Prophylaxis | Outcome |
| --- | --- | --- | --- | --- | --- | --- | --- |
| 1 | 70s | F | 3 | Mechanical | Acute PE | SQ Heparin 5000 U TID | Discharge |
| 2 | 60s | M | 9 | Mechanical | Ischemic stroke (left middle cerebral artery) | SQ Heparin 5000 U TID | Discharge |
|  |  |  | 19 | Mechanical | LE DVT (right femoral vein, iliac vein, and popliteal vein) | SQ Heparin 7500 U TID |  |
| 3 | 50s | M | 16 | Mechanical | Ischemic stroke (left posterior cerebral artery) | SQ Heparin 5000 U TID | Death |
| 4 | 50s | M | 27 | Mechanical | Ischemic stroke (both supra- and infra-tentorial foci, suggesting an embolic source) | SQ Heparin 7500 U TID | Remains in hospital |
| 5 | 30s | M | 2 | Mechanical | Bilateral LE DVT (right popliteal vein, left gastrocnemius vein) | Enoxaparin 40 mg daily | Discharge |
| 6 | 40s | M | 2 | Mechanical | Bilateral LE DVT (bilateral common femoral veins and popliteal veins) | SQ Heparin 5000 U TID | Discharge |
| 7 | 40s | M | 36 | Mechanical | Acute PE (segmental and subsegmental) | Heparin gtt 1400 U/hour | Death |
| 8 | 40s | F | 1 | Nasal cannula | Acute PE (segmental and subsegmental) | None | Discharge |
| 9 | 50s | M | 1 | Room air | Acute PE (segmental and subsegmental) | None | Discharge |
| 10 | 60s | M | 5 | Mechanical | UE DVT (right UE DVT) | Enoxaparin 40 mg daily | Discharge |
| 11 | 70s | M | 9 | Mechanical | Acute PE (segmental) | Heparin gtt 850 U/hour | Discharge |

F=female; M=male; SQ=subcutaneous; gtt=continuous IV heparin; LE=lower extremity; UE=upper extremity; DVT=deep vein thrombosis; PE=pulmonary embolism; u=units; TID=three times daily

To better understand the extent to which patients diagnosed with thrombotic events differed from other patients hospitalized with COVID-19, we identified a matched cohort of 33 patients hospitalized with COVID-19 over the same month (**Table 1**). The control cohort was identical for age and sex. The cohort was similar in terms of comorbidities and ultimate outcome (18% died in the thrombosis group and 21% in the control group). For all 44 patients, we were able to access a blood sample collected during their hospitalization. As it relates to diagnosis of the first thrombotic event for each patient, six blood samples were banked within three days of event diagnosis. Three were banked earlier (5, 7, and 12 days prior to event diagnosis) and two later (both at 6 days). As compared with the control group, patients with a thrombotic event demonstrated significantly higher levels of calprotectin, a marker of neutrophil activation (**Figure 1A**). Similarly, three different markers of NETs (cell-free DNA, myeloperoxidase-DNA complexes, and citrullinated histone H3) were also markedly elevated in the thrombosis group as compared with the matched controls (**Figure 1B-D**).

**Figure 1:**
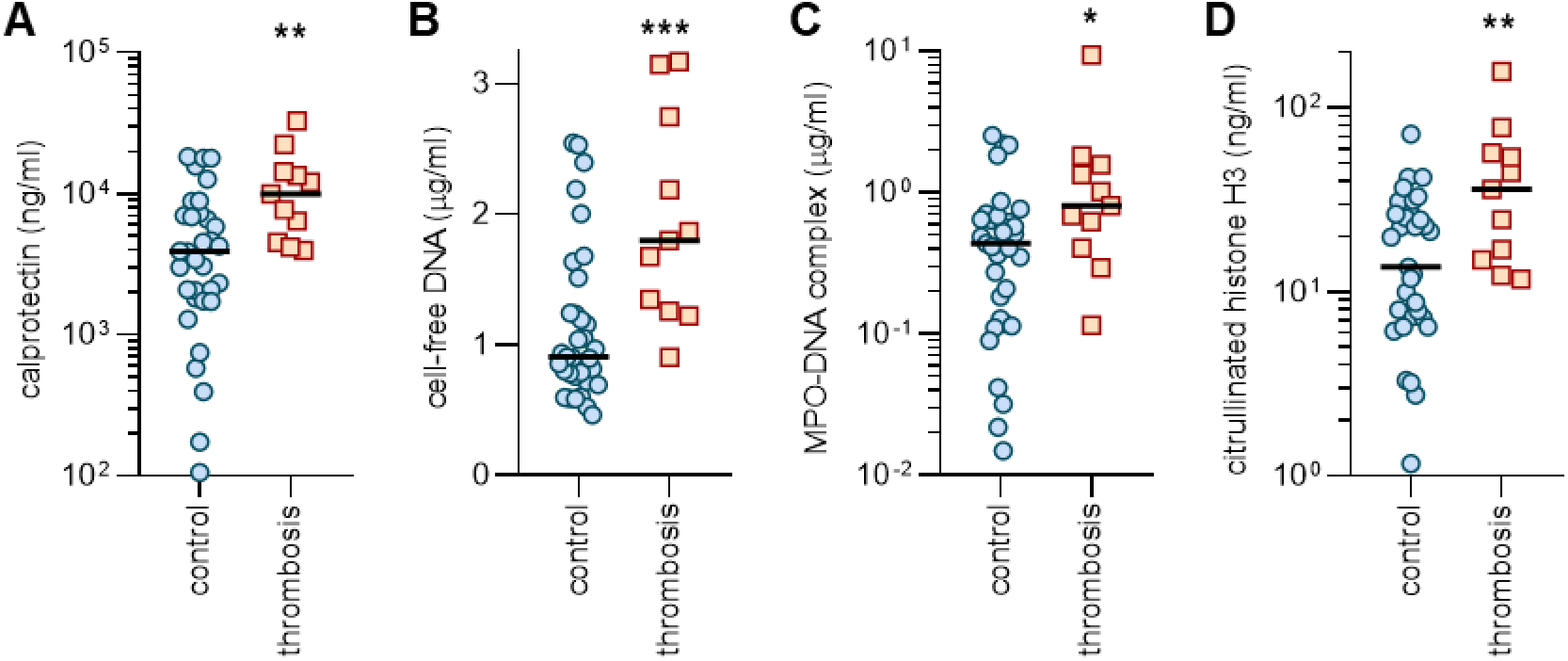
Elevated levels of NETs in the blood of COVID-19 patients diagnosed with a thrombotic event, as compared with matched controls. Serum was tested for calprotectin (**A**), cell-free DNA (**B**), myeloperoxidase-DNA complexes (**C**), and citrullinated histone H3 (D). N=33 for the control group and n=11 for the thrombosis group. Comparisons were by Mann-Whitney test; *p<0.05, **p<0.01, and ***p<0.001.

We then turned our attention to other clinical biomarkers that might associate with a thrombotic event. The thrombosis group had higher levels of peak D-dimer (**Figure 2A**), but not troponin (**Figure 2B**). Peak CRP and ferritin were also modestly higher in the thrombosis group (**Figure 2C-2D**). Interestingly, peak neutrophil levels did not differ between groups, but platelets were significantly higher in patients who developed thrombosis (**Figure 2E-F**). Finally, we asked whether there was an association between blood markers of neutrophil activation (such as calprotectin and cell-free DNA) and D-dimer within this cohort of COVID-19 patients (n=44). Despite the small number of patients, there were strong correlations between peak D-dimer and both calprotectin and cell free DNA (**Figure 3A-B**).

**Figure 2:**
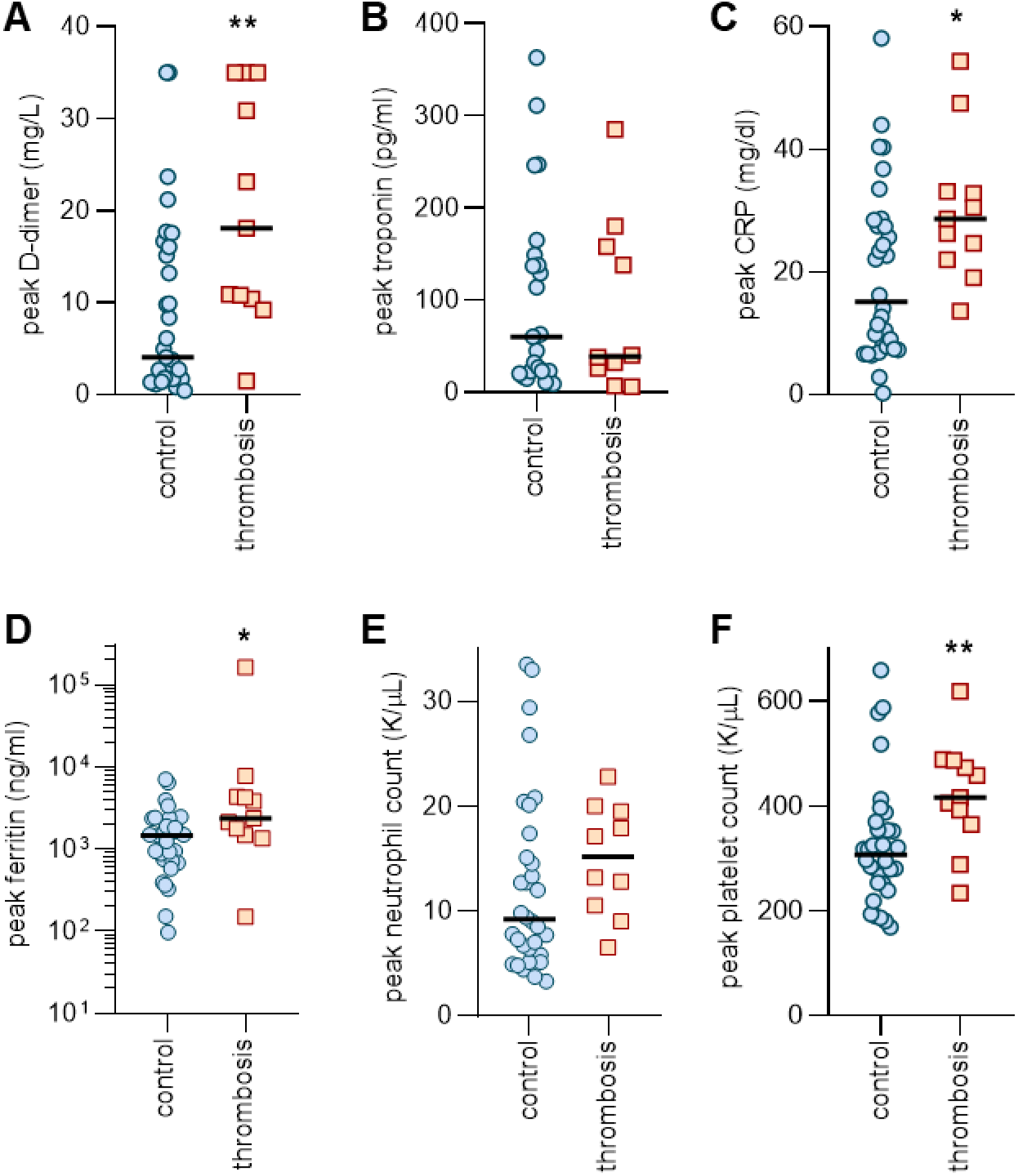
Association between peak levels of clinical biomarkers and diagnosis of a thrombotic event. Clinical testing is reported for D-dimer (**A**), troponin (**B**), C-reactive protein (**C**), ferritin (**D**), absolute neutrophil count (**E**), and absolute platelet count (**F**). N=33 for the control group and n=11 for the thrombosis group. Comparisons were by Mann-Whitney test; *p<0.05 and **p<0.01. Comparisons for peak troponin and peak neutrophil count were not statistically significant.

**Figure 3:**
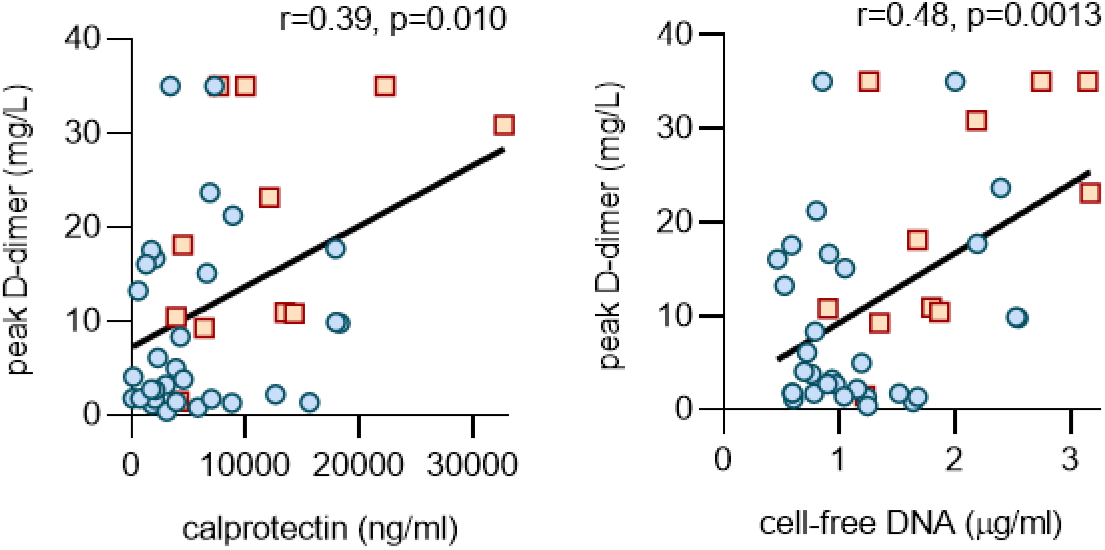
Correlation between neutrophil activation markers and D-dimer. Calprotectin (**A**) and cell-free DNA (**B**) were compared to peak D-dimer levels. Data were analyzed by Pearson’s method.

## DISCUSSION

Hyperactivity of the coagulation system is a common finding of severe COVID-19 [20]. Indeed, many patients have a profile to suggest a prothrombotic diathesis including high levels of fibrin degradation products (D-dimer), elevated fibrinogen levels, and low antithrombin levels [20, 21]. We recently characterized the blood of 50 patients hospitalized with COVID-19 and found significantly elevated levels of NETs as compared with healthy controls [18]. Here, we extend that analysis by demonstrating particularly high levels of NETs in a subgroup of COVID-19 patients diagnosed with a thrombotic event. Given the known link between NETs and thrombosis in many inflammatory conditions, these data suggest that the role of NETs in COVID-19-associated thrombophilia warrants systematic investigation.

Intravascular NET release is responsible for initiation and accretion of thrombotic events in arteries, veins, and microvessels, where thrombotic disease can drive end-organ damage in lungs, heart, kidneys, and other organs[22, 23]. Mechanistically, DNA in NETs may directly activate the extrinsic pathway of coagulation[24], while NETs also present tissue factor to initiate the intrinsic pathway[25]. Serine proteases in NETs such as neutrophil elastase release brakes on coagulation by proteolyzing various tissue factor pathway inhibitors [26]. Bidirectional interplay between NETs and platelets might also be critical for COVID-19-associated thrombosis as has been characterized in a variety of disease models [23, 24].

Approaches to combatting NETs [27-29] include the dismantling of NETs with deoxyribonucleases and strategies that prevent initiation of NET release such as neutrophil elastase inhibitors and peptidylarginine deiminase 4 inhibitors. As we await definitive antiviral and immunologic solutions to the current pandemic, we posit that anti-neutrophil therapies may be part of a personalized strategy for some individuals affected by COVID-19. Furthermore, those patients with hyperactive neutrophils may be at particularly high risk for thrombotic events and might therefore benefit from more aggressive anticoagulation while hospitalized.

## Data Availability

Once published in a peer-reviewed journal, all primary data will be available upon request to the corresponding authors.

## ACKNOWLEDGEMENTS

The work was supported by a COVID-19 Cardiovascular Impact Research Ignitor Grant from the Michigan Medicine Frankel Cardiovascular Center as well as by the A. Alfred Taubman Medical Research Institute. YZ was supported by career development grants from the Rheumatology Research Foundation and APS ACTION. JAM was partially supported by the VA Healthcare System. JSK was supported by grants from the NIH (R01HL115138), Lupus Research Alliance, and Burroughs Wellcome Fund. YK was supported by the NIH (K08HL131993, R01HL150392), Falk Medical Research Trust Catalyst Award, and the JOBST-American Venous Forum Award. The authors also thank all members of the “NETwork to Target Neutrophils in COVID-19” for their helpful advice and encouragement.

## AUTHORSHIP

YZ, MZ, SY, KG, JM, HS, WW, and SPL conducted experiments and analyzed data. YZ, MZ, NLL, YK, and JSK conceived the study and analyzed data. All authors participated in writing the manuscript and gave approval before submission.

## Competing interests

The authors have no financial conflicts to disclose.

## Notes

### Competing Interest Statement

The authors have declared no competing interest.

### Author Declarations

This study complied with all relevant ethical regulations, and was approved by the University of Michigan Institutional Review Board (HUM00179409).

